# Clinical Epistemic Reopening: When Patient Reality Makes Medical Framing Reconsiderable

**DOI:** 10.64898/2026.10.10.26365203

**Authors:** Keiko Yoshimura, Sae Fujioka, Paul Matthew Lewis, Mayuko Hirata, Misato Tani, Kohei Akazawa, Norio Kohno

## Abstract

Modern clinical medicine depends on reducing complex human realities into forms that support knowledge, judgment, and action. Such reduction is indispensable to scientific medicine, but necessarily selective. A medical framing may therefore remain well warranted within medicine while becoming insufficient in relation to the reality of a particular patient. What happens clinically when the adequacy of such a framing itself becomes uncertain remains insufficiently understood.

We conducted an abductive, theory-generating qualitative study of 44 patients who attended a supplementary Dialogue-Based Outpatient Clinic within a breast oncology service in Japan between 2021 and 2023, comprising 111 consultations. The clinic had no predefined clinical agenda, allowing patients to raise concerns that mattered to them. Through iterative comparison within and across cases, we examined how patients’ concerns were understood and responded to clinically.

Across cases, a recurrent clinical process became apparent when patient reality called the adequacy of existing medical framing into question. Exploration of patients’ concerns could expose a consequential mismatch between their reality and the framing through which medicine understands and represents clinical reality. Such mismatches prompted inquiry into medical framing to identify what required reopening. Reopening occurred when the identified framing, previously treated as provisionally settled, became reconsiderable in relation to the patient’s reality. Reconsideration involved examining what had been reopened and exploring clinical responses through dialogue between clinician and patient, followed by a clinical response and provisional closure. We describe this recurrent process as Clinical Epistemic Reopening (CER). CER identifies a clinical process through which the framing produced by medicine’s necessary selectivity in making complex reality clinically knowable and actionable can itself become reconsiderable when patient reality reveals its limits. CER can thereby reverse the usual direction of clinical scrutiny: aspects of medicine that ordinarily function as means of understanding and acting upon the patient can themselves become objects of clinical inquiry. CER thus allows medicine to remain open to patient reality beyond the limits of its existing framings.

**Highlights:** Patient reality can challenge the adequacy of warranted medical framing. Reopening shifts framing’s epistemic status from settled to reconsiderable. In CER, aspects of medicine itself become objects of clinical inquiry.

CER offers a way for medicine to remain open to patient reality beyond the limits of its existing framings.

## 1. Introduction

Modern medicine depends on reducing complex human phenomena into forms that can be observed, classified, compared, and acted upon. Diagnostic categories, clinical measurements, medical images, prognostic models, randomized trials, and practice guidelines make possible forms of knowledge and intervention that would otherwise be unattainable. Reduction is therefore one of the conditions of its effectiveness (Montgomery, 2005).

Yet reduction necessarily entails selection. Some aspects of reality become visible within a medical frame, while others become less visible or remain outside it. Through reduction, classification, and prioritization, medicine constructs workable representations of complex realities and frames clinical problems in ways that provisionally settle what counts as the problem, what evidence is relevant, what outcomes matter, and what actions are justified (Fleck, 1981; Hanson, 1958; Montgomery, 2005). Such provisional closure is indispensable to clinical practice. Clinicians cannot indefinitely suspend judgment while attempting to apprehend the totality of a patient’s reality. Diagnoses must be made, investigations selected, recommendations offered, and decisions taken. Yet the same selective framing through which reality is made medically intelligible may become insufficient in relation to a particular patient when consequential aspects of that patient’s reality fall outside, or come into tension with, what that framing makes visible and knowable. The question, therefore, is not whether medicine should reduce or frame reality, but how it can remain responsive when the adequacy of an otherwise warranted medical framing becomes uncertain in relation to the reality of the particular patient.

The limits of medical framing have been approached across several intellectual traditions, including through critical examinations of medicine itself. Philosophy of science has examined how observation, evidence, and explanation are shaped by conceptual and theoretical frameworks (Fleck, 1981; Hanson, 1958; Kuhn, 1970). Medical anthropology has distinguished biomedical accounts of disease from patients’ experiences of illness and examined how differing understandings of illness shape clinical encounters (Good, 1994; Kleinman et al., 1978; Kleinman, 1988). Sociology and the history of medicine have examined how medical knowledge, categories, and practices are shaped by social and historical conditions, and how medicine in turn shapes the understanding and experience of health, illness, and disease (Conrad, 1992; Rosenberg, 1989, 2002, 2003), while medical humanities have explored dimensions of meaning, embodiment, and suffering, as well as critically examined the assumptions and practices of medicine itself (Charon, 2001; Viney et al., 2015). Work in clinical epistemology has examined how patients’ experiential knowledge may be marginalized within healthcare and how the epistemic authority and limits of medical knowledge bear on clinical practice (Carel and Kidd, 2014; Kidd and Carel, 2017; Schwab, 2012). Across these traditions lies a common recognition: medically useful accounts of reality do not exhaust the reality of the person to whom they are applied.

Related concerns have also been addressed through several traditions in clinical practice. The biopsychosocial model, patient-centred care, narrative medicine, shared decision-making, and work in medical anthropology have, in different ways, sought to bring patients’ experiences, circumstances, values, meanings, and perspectives more fully into clinical care (Charon, 2001; Clayman et al., 2017; Elwyn et al., 2012; Engel, 1977; Epstein and Street, 2007; Kleinman et al., 1978; Montori et al., 2023; Stewart et al., 1995). These approaches have substantially broadened what medicine takes into account and how clinicians understand the persons they care for.

An important analytical distinction can be made between bringing more aspects of the patient’s reality into clinical consideration and reconsidering the medical framing itself through which that reality is made clinically intelligible and actionable. A clinician may attend carefully to family relationships, life history, work, identity, values, preferences, and the patient’s own explanations without necessarily calling into question the framing through which the clinical problem has been constituted. In many situations, this is entirely appropriate. Additional information, better communication, reconciliation of differing understandings, or fuller incorporation of the patient’s perspective may be sufficient.

At other times, however, patient reality may become relevant not only to what is considered within a medical frame, but to the adequacy of the frame itself. Something that emerges through the clinical encounter may reveal that the difficulty lies not simply in what has yet to be included, but in how the clinical problem has been framed. At such moments, what changes is not necessarily the medical knowledge itself, but the epistemic status of the framing through which that knowledge has been brought to bear: what has been provisionally settled may become reconsiderable. The clinical task may then involve not only responding to the patient within an existing medical framing, but bringing that framing itself into renewed clinical consideration.

What remains insufficiently understood is what happens clinically when the adequacy of an otherwise warranted medical framing itself becomes uncertain in relation to a particular patient’s reality, and how the framing may then enter clinical inquiry and renewed consideration.

Through an abductive, theory-generating analysis of consultations conducted in a Dialogue-Based Outpatient Clinic embedded within a breast oncology service in Japan, a recurrent clinical process became apparent. In this process, engagement with patients’ concerns could expose a consequential mismatch between some aspect of patient reality and the medical framing through which the clinical problem had been understood, prompting inquiry into that framing and, where warranted, its selective reopening and reconsideration. We term this process Clinical Epistemic Reopening (CER). This article develops the concept empirically from these clinical encounters.

## 2. Methods

### 2.1. Study design

We conducted an abductive, theory-generating qualitative study of consultations in a Dialogue-Based Outpatient Clinic. The study examined what became clinically visible when consultations began from concerns that mattered to patients and the clinician sought to respond to those concerns as part of clinical care. The clinic did not prescribe a specific consultation method in advance, allowing the clinical approach to be guided by the concerns raised by each patient. An abductive approach allowed iterative movement between the clinical material and emerging conceptual interpretations as recurrent patterns and unexpected findings were examined across cases.

### 2.2. Study setting

The Dialogue-Based Outpatient Clinic was established within a breast oncology service as a complementary clinical setting alongside routine oncology care. Patients attended either on their own initiative or following a suggestion from a healthcare professional. The timing and agenda of consultations were not predefined; patients could seek consultation when they wished and determine what they wished to discuss.

The clinic developed from a longstanding clinical concern that, in making complex realities clinically manageable, medicine might leave consequential aspects of individual patients’ realities insufficiently addressed within routine clinical practice. The clinic was established on the premise that biomedical understanding, while essential, does not encompass the whole of a patient’s lived reality. Providing a setting in which patients could raise concerns freely was intended both to complement routine care and to allow such difficulties, if present, to become more visible. Consultations sought to understand patients’ concerns without requiring them to conform to a predefined medical perspective, and to consider the clinical situation together from that starting point.

The clinic was not, however, designed to implement or test any specific theoretical framework, predefined categories, or method. Consultations remained part of clinical care rather than research encounters, with biomedical assessment and clinical judgment remaining integral throughout.

### 2.3. Participants

The study included all 44 patients who attended the Dialogue-Based Outpatient Clinic between 2021 and 2023, across 111 consultations. Patients had breast cancer or were undergoing evaluation for suspected breast cancer at the time of their first visit to the clinic. No patients who attended during the study period were excluded from the analysis.

### 2.4. Data collection

Data consisted of routine electronic clinical records and, when the content of the consultation could not be adequately captured in the electronic clinical record, supplementary records prepared by the consulting physician, both documented during or after the consultations. These records documented the concerns raised by patients, relevant clinical context, aspects of the dialogue, clinical considerations, and subsequent responses or actions. Because the consultations were conducted as part of routine clinical practice rather than as research interviews, they were not audio-recorded, and verbatim transcripts were not produced.

### 2.5. Data analysis

Analysis proceeded through repeated reading of the consultation records and iterative comparison within individual cases and across cases, consistent with an abductive approach to theory construction (Timmermans and Tavory, 2012). Attention was initially directed to what patients brought to the consultations, how their concerns were understood and responded to clinically, and what patterns and differences became apparent across cases.

During the early consultations at the Dialogue-Based Outpatient Clinic, the consulting physician recognized a distinction between concerns that could be addressed within routine medical care and those that appeared to require consideration beyond its usual scope. This initial clinical observation was subsequently examined and refined through iterative analysis across all 44 patients.

Through this comparison, the initial clinical distinction was further classified between cases in which patients’ concerns could be addressed while the existing medical framing remained adequate and cases in which aspects of patients’ realities called the adequacy of that framing into question. In the latter cases, analysis increasingly focused on how engagement with the patient’s concerns brought the medical framing through which the clinical problem had been understood into question, and how that framing was subsequently reconsidered. This shifted the analytic focus from the content of patients’ concerns alone toward how the adequacy of medical framing itself came into question in relation to the patient’s reality.

Although patients’ concerns and the medical framings brought into question varied across cases, continued comparison revealed a recurrent process in how those framings came into question and were reconsidered. This recurrent process was progressively refined through further iterative comparison, while contrasting cases in which existing framing remained adequate were retained throughout the analysis. The analytic distinctions of consequential mismatch, inquiry into medical framing, reopening, reconsideration, clinical response, and provisional closure were developed through this process. Although the reconsideration described here occurred within the clinical encounters, its recurrent conceptual structure as CER was identified retrospectively through this analysis. The consulting physician led the primary case-level analysis, while emerging interpretations and the developing conceptual structure were further examined through critical discussion within the research team.

### 2.6. Researcher positioning and reflexivity

The physician who established the Dialogue-Based Outpatient Clinic and conducted the consultations had more than 20 years of clinical experience in breast oncology and had also engaged with medical anthropology and related interdisciplinary scholarship.

This dual role provided close access to the clinical reasoning and clinical context of the consultations, while also requiring reflexive attention to how the physician’s clinical perspective might influence the interpretation of the encounters. Reflexivity was maintained through repeated return to the consultation records, consideration of alternative interpretations, including whether observed patterns could be adequately accounted for by existing clinical concepts, and critical discussion within the research team.

### 2.7. Ethical considerations

The study was approved by the institutional ethics committee of Osaka Saiseikai Nakatsu Hospital (approval number 2026-60). Participation was managed through an opt-out process in accordance with the approved protocol. Patient privacy was protected throughout the study, and identifying information was minimized in the presentation of cases.

## 3. Results

### 3.1. From Patient Concerns to Medical Framing

Analysis of the concerns patients brought to the consultations showed wide variation in their content and in how they became clinically consequential. Participant characteristics are summarized in Table 1. Across this diversity, however, a distinction became apparent. Some concerns could be addressed while the existing medical framing remained adequate, whereas in others, aspects of patients’ realities became consequential in ways that brought the adequacy of that framing itself into question. Comparison across the full set of cases revealed two broad patterns.

**Table 1.** Characteristics of study participants.

| Characteristic | Total (n = 44) | Existing medical framing remained adequate (n = 13) | Consequential mismatch with existing medical framing identified (n = 31) |
| --- | --- | --- | --- |
| **Age, years** |  |  |  |
| 30–39 | 3 (7%) | 2 (15%) | 1 (3%) |
| 40–49 | 8 (18%) | 4 (31%) | 4 (13%) |
| 50–59 | 11 (25%) | 1 (8%) | 10 (32%) |
| 60–69 | 13 (30%) | 3 (23%) | 10 (32%) |
| 70–79 | 4 (9%) | 1 (8%) | 3 (10%) |
| 80–89 | 5 (11%) | 2 (15%) | 3 (10%) |
| **Disease status** |  |  |  |
| Primary breast cancer | 40 (91%) | 13 (100%) | 27 (87%) |
| Recurrent breast cancer | 4 (9%) | 0 (0%) | 4 (13%) |
| **Treatment status at first Dialogue-Based Outpatient Clinic consultation** |  |  |  |
| Before diagnosis | 1 (2%) | 0 (0%) | 1 (3%) |
| Before surgery | 8 (18%) | 4 (31%) | 4 (13%) |
| Before chemotherapy | 1 (2%) | 0 (0%) | 1 (3%) |
| Before radiotherapy | 1 (2%) | 0 (0%) | 1 (3%) |
| Receiving endocrine therapy | 20 (45%) | 8 (62%) | 12 (39%) |
| Receiving chemotherapy | 6 (14%) | 1 (8%) | 5 (16%) |
| Receiving endocrine therapy plus chemotherapy | 1 (2%) | 0 (0%) | 1 (3%) |
| Observation | 6 (14%) | 0 (0%) | 6 (19%) |

*Data are presented as n (%). Percentages are calculated within each analytic group and may not total 100% because of rounding. One patient first attended the Dialogue-Based Outpatient Clinic before a definitive diagnosis of breast cancer and was subsequently diagnosed with primary breast cancer.*

In 13 patients, the existing medical framing remained adequate for addressing the concern. The concerns could be addressed through information or explanation, diagnostic evaluation, or other clinical responses within the existing framing. Patients’ circumstances could likewise be considered within that framing, without requiring examination of the adequacy of the framing itself (Supplementary Table S1).

In the remaining 31 patients, as their concerns were explored, aspects of patients’ realities emerged as clinically relevant in relation to the existing medical framing. A mismatch became apparent between some aspect of the patient’s reality and the framing through which the clinical problem had been understood. This did not necessarily indicate that the medical knowledge or judgment involved was incorrect. Rather, medically warranted knowledge and judgments could remain valid while the framing through which they were brought to bear on the particular patient proved insufficient in relation to that patient’s reality. We refer to this form of mismatch as a consequential mismatch.

Consequential mismatches took different forms across the 31 patients. Engaging with these mismatches prompted inquiry into the medical framing through which the clinical situation had been understood. Patient reality therefore mattered not simply as additional information to be incorporated into care, but by bringing medicine’s own way of understanding and responding to the clinical situation into inquiry. The following section examines how such mismatches became apparent and how they shaped subsequent clinical response in representative encounters.

### 3.2. From Consequential Mismatch to Clinical Response

Consequential mismatches became apparent in different, sometimes overlapping forms across the 31 patients. Six cases were selected for detailed presentation to illustrate the diversity of clinical situations and of the aspects of medical framing brought into question.

In some cases, the mismatch became apparent at the limits of what medicine could establish or legitimately conclude. A patient with intermittent abdominal pain had undergone repeated medical evaluation at several hospitals without an identifiable cause. Yet the continuing pain continued to matter to the patient. She remained troubled by the pain and by being told that nothing was wrong, and came to the Dialogue-Based Outpatient Clinic seeking guidance on how to understand and respond to it. The negative evaluation remained clinically meaningful: it provided reassurance that no condition requiring current medical intervention had been identified, but it had not resolved the question of how her continuing pain should be understood. What became reconsiderable was the reach of medical investigation itself. Medical investigations do not apprehend a clinical reality in its entirety; they examine particular aspects of it through specific methods and technologies. What they can establish is therefore bounded by what those methods can capture. These limits were discussed explicitly with the patient, including that not all mechanisms underlying abdominal pain are fully understood.

The negative findings could therefore be understood within the reach of what the investigations could establish, without negating the continuing pain. What had been established, what remained unknown, and what could reasonably follow from that uncertainty were considered together with the patient. Through this dialogue, continued observation came to make sense to her as a reasonable way forward, with the understanding that she could return if her symptoms changed or further concerns arose. The pain subsequently subsided without further intervention.

In another case, aspects of medical framing and intervention themselves had become implicated in the patient’s concern. A woman who underwent mastectomy after neoadjuvant chemotherapy achieved a pathological complete response (pCR), a favorable oncological outcome in which no residual cancer was identified in the resected specimen. Nevertheless, she experienced profound distress both from the loss of her breast and from learning that no residual cancer had ultimately been found in the breast that had been removed. While the treatment was regarded as successful, her continuing distress had been framed primarily as a psychiatric problem, which further intensified her suffering. Feeling that this framing did not address what was troubling her, she sought consultation with a breast clinician at the clinic. She described her experience as one in which her disease had been treated, but she herself had not been cared for as a person.

Pathological complete response remained a genuine oncological success. For the patient, however, that very success intensified the tension between oncological success and the irreversible loss of her breast. She was therefore left with a question that neither the oncological account of treatment success nor the psychiatric framing of her continuing distress could adequately answer: Why had losing her breast been medically necessary?

What became reconsiderable was neither the validity of the oncological outcome nor the validity of the medical reasoning that had led to treatment. Rather, the question was whether, in responding to the consequences of that treatment, medicine’s own reasoning―itself situated within the knowledge and possibilities of its time―and its account of “treatment success” should remain outside clinical scrutiny. Medicine no longer functioned only as the framework through which the patient’s experience was interpreted; medicine itself—its reasoning, possibilities, limitations, and historical situatedness—became available for examination within the clinical encounter.

Clinician and patient examined this together, including why mastectomy had been medically warranted at the time despite the subsequent finding of pCR. Their inquiry extended beyond the individual treatment decision to how breast cancer treatment itself had shifted historically alongside changes in medical knowledge, reasoning, diagnostic capabilities, and therapeutic possibilities, situating medicine itself within its historical conditions of knowledge and practice. Through this reconsideration, she came to understand that her treatment had been undertaken in her interests within medical knowledge and possibilities that were themselves historically situated.

This allowed pathological complete response to remain a valid oncological account of treatment success while the irreversible loss of her breast could also remain consequential to what that treatment meant in her life. Neither had to invalidate the other. Her distress no longer needed to be located solely within her as a psychiatric problem. Medicine itself could remain implicated in what had occurred and participate in responding to its consequences.

Through this understanding, she gradually became able to accept the loss of her breast and came to regard both her treating physician and the medical care she had received with renewed warmth. At the same time, the clinical question expanded beyond how her distress might be alleviated to what medicine could still do in response to an irreversible loss arising from medical treatment.

Within this reconsidered understanding, breast reconstruction emerged not simply as another procedure, but as a clinically meaningful possibility in relation to the loss she had experienced and how she wished to live thereafter. She chose reconstruction and subsequently underwent reconstructive surgery.

In still other encounters, the patient’s reality made the purpose organizing medical care itself visible as something that could no longer simply be assumed. An older woman living independently with little family or social support was found on CT to have a small pulmonary lesion of uncertain malignant potential. Because malignancy could not be excluded, her treating physician had recommended surgical resection to establish a diagnosis and, if malignant, provide treatment. She subsequently sought consultation because she was concerned that surgery itself might disrupt the independent daily life she was managing on her own.

In the consultation, the uncertainties on both sides were considered. Resection could establish a diagnosis and potentially enable treatment, while foregoing surgery carried the possibility of malignant progression. At the same time, if the lesion proved benign, surgery would retrospectively have exposed her to an intervention from which she derived no oncological benefit. Nor was it certain that surgery would disrupt her independent life; she might recover and continue living much as before, although disruption remained a possibility that mattered greatly to her. The clinician and patient therefore considered together that medicine could not determine in advance which course would ultimately prove least harmful.

As these possibilities were considered together, a consequential mismatch became visible. It lay not simply between a recommended intervention and the patient’s preference, but between the purpose ordinarily organizing the proposed medical course and the purpose medicine was to serve in her present situation. The clinician explained that if prolonging life was the primary purpose medicine was to serve, pursuing diagnosis and treatment would be the medically more coherent course. The more fundamental question was therefore made explicit: what was medicine to be used for in her situation? If prolongation of life was not to be its primary purpose, care would need to be reconsidered in relation to the purpose medicine was instead to serve.

What became reconsiderable was therefore not merely which intervention she preferred, but the purpose according to which medical knowledge and intervention should be organized in her care. Diagnosis, treatment of possible disease, and life prolongation remained medically warranted aims; what was no longer taken for granted was that they should, by virtue of their medical warrant alone, determine what medicine was for in her situation. Through this reconsideration, that purpose became clearer: not primarily to pursue prolongation of life, but to preserve, as far as possible, the independent life she was presently able to sustain. The clinical question consequently shifted from whether the pulmonary lesion should be surgically diagnosed to how medicine should be used in relation to the purpose that had emerged through reconsideration.

She chose surveillance, with the possibility of reconsideration if the lesion, her circumstances, or the purpose medicine was to serve changed over time. The lesion remained stable during subsequent follow-up and was ultimately regarded as benign.

The same issue could arise at the other end of the clinical course, where a change in the purpose of care made the meaning of continued medical involvement itself uncertain. A patient in her 40s with metastatic disease subsequently sought consultation at the clinic after telling her treating physician that she did not wish to pursue further disease-directed treatment. In the consultation, after carefully exploring with her the basis of her decision and confirming that she understood her condition, available treatment options, and the implications of foregoing further disease-directed treatment, the question of treatment choice was largely settled. Yet she had come to the clinic with a concern that remained: she did not seek another intervention, but wanted a physician who understood her medical condition and how she wished to approach her life and death to continue to be present as her disease progressed.

Her request made visible an assumption implicit in the usual course of care: that when altering the course of disease was no longer the purpose of medical involvement, the physician’s role would correspondingly recede. The consequential mismatch therefore concerned not which treatment she should receive but what continued medical involvement was for once disease-directed treatment was no longer the purpose organizing care.

What became reconsiderable was the purpose of continued medical involvement and, with it, the physician’s role. Medical knowledge of her condition remained relevant even when it was no longer being used primarily to guide disease-directed intervention. The more fundamental question was whether such knowledge and the physician’s involvement had clinical meaning only insofar as they served intervention, or whether they could be organized toward a different purpose. In her situation, continued medical involvement could instead allow a physician who understood both her disease and the depth with which she had considered and accepted the possibility of death in foregoing further treatment to remain medically present as her condition evolved.

Under this reconsidered purpose, the physician agreed to continue seeing her, with medical knowledge and clinical presence organized not primarily toward altering the disease course, but toward accompanying her through its progression while remaining attentive to both her condition and the way she had chosen to approach her life and death.

Some encounters exposed a different kind of mismatch: between what the patient sought medical knowledge to determine and what that knowledge could legitimately determine for an individual patient. A patient with bilateral breast cancer had been presented by her treating physician with two surgical approaches: breast-conserving surgery followed by radiotherapy to both breasts, or bilateral mastectomy. Both were medically reasonable, and she sought consultation at the clinic because medical explanations had not resolved which she should choose. There, she repeatedly sought confirmation that one was the medically “right” decision. Medical knowledge could compare the two approaches, their expected benefits and harms, and their oncological implications. Both remained medically warranted, however, and further comparison could not determine which was the “right” choice for this individual.

Her repeated search for a medically correct answer made visible an assumption that had been organizing the question itself: that sufficient medical knowledge should be able to determine which choice she ought to make. When both options remained medically warranted, the adequacy of that assumption became uncertain. What became reconsiderable was therefore not the validity or sufficiency of the medical information itself, but the scope of what medical knowledge could legitimately determine for an individual patient. The clinician and patient considered this boundary explicitly. Rather than extending medical authority beyond what the available knowledge could determine, they explored what might reasonably ground her decision among considerations that had remained outside the medical framing of the choice. Among these, she came to place particular value on making a choice she believed she would not regret and ultimately chose bilateral mastectomy. The clinical response therefore did not arise from resolving the uncertainty, but from making the limit of medical determination itself part of the clinical reasoning through which the decision could be made.

A related boundary became visible in another patient considering whether to continue endocrine therapy beyond the duration usually recommended for early breast cancer. A close friend of hers had died of breast cancer despite receiving treatment in accordance with medical evidence. This experience had brought into sharp focus for her the relationship between what evidence could predict and what would actually happen to an individual person. Here, the problem was not the validity of the available medical evidence, but the necessarily probabilistic nature of what it could establish. Evidence could inform the expected benefits and harms of extended treatment at the population level, but it could not determine in advance whether continued treatment would ultimately benefit this particular patient. The consequential mismatch therefore lay between what valid medical knowledge could establish about expected outcomes in a population and what the patient sought it to determine about her own future.

In reconsidering this mismatch, the validity of the evidence itself was not called into question. Her friend’s experience was likewise not treated as evidence against population-level knowledge. Rather, it brought the limits of such knowledge for individual prediction into clinical focus. What became reconsiderable was the assumption that valid population-level knowledge should be able to yield a medically correct answer for the individual patient. Once that limit was made explicit, the question was no longer how to obtain enough medical knowledge to eliminate uncertainty, but how that knowledge should inform a decision that necessarily remained underdetermined by it. The clinician and patient could then consider continued endocrine therapy, in light of what the evidence could and could not determine for her individually, without treating the remaining uncertainty as a deficit that further medical knowledge could necessarily resolve.

Across these two encounters, the boundary that became visible was not a failure of medical knowledge, but a limit in what valid medical knowledge could legitimately determine for an individual patient. Making that boundary itself reconsiderable allowed clinical reasoning to proceed without either dismissing medical knowledge or extending its authority beyond what it could support.

Viewed across the encounters described above, consequential mismatches became apparent at different points in the relation between medical framing and the patient’s reality. Following what had become clinically salient did not necessarily lead only to the particular framing in which the mismatch had first appeared; in some encounters, it brought a related or more fundamental medical framing into question. This could include how medical findings or evidence were interpreted, how the clinical problem itself was framed, how medicine’s own reasoning and limitations were understood, how interventions were warranted, what purposes care should serve, what the clinician’s role could entail, or what medical knowledge could legitimately determine. Across these encounters, a further pattern became visible. Medical framing, which ordinarily functioned as a means of understanding the patient, determining what should be done, and acting upon the patient, could itself become an object of clinical inquiry when the patient’s reality made its adequacy in the particular situation uncertain. Clinical scrutiny could therefore change direction: rather than only asking how the patient’s difficulty should be understood within an existing medical framing, the encounter could also bring into question the framing through which that difficulty had become clinically intelligible in the first place. What had previously functioned as settled within medical reasoning could thereby become reconsiderable.

Comparison across these heterogeneous encounters revealed a recurrent movement from consequential mismatch to the reopening of what had previously been settled within medical reasoning. The recurrent process through which this occurred is described in the following section.

### 3.3. The Recurrent Process of Clinical Epistemic Reopening

Across these encounters, the recurrent movement described above could be characterized as a clinical process with a discernible structure. The recurrent process began when a consequential mismatch between the patient’s reality and existing medical framing prompted inquiry into that framing. This inquiry did not necessarily remain focused on the framing in which the mismatch first appeared; it could extend to a related or more fundamental framing. Its depth was not fixed in advance; inquiry proceeded from the consequential mismatch until what required reopening in relation to that patient became identifiable. When the adequacy of what had previously been settled could no longer be assumed in the particular situation, what had been settled became reconsiderable. We refer to this change as reopening.

Reopening did not imply that the existing medical framing had been mistaken, nor did it require medical reasoning as a whole to be reopened. Medical knowledge, judgments, and framing that remained warranted and adequate for the particular patient could remain settled. What changed was the epistemic status of what the encounter had placed in question. Across cases, what became reconsiderable was not limited to the application of medical knowledge to individual circumstances. Medicine’s own ways of knowing, representing, intervening, assigning authority, defining clinical purpose, and assuming responsibility could themselves become objects of clinical reconsideration.

Reconsideration involved working with what had been reopened in relation to the patient’s reality. The reopened framing could be made explicit and brought into dialogue in accessible terms. Clinician and patient could consider what medicine could establish, what remained uncertain, what an intervention was for, what purposes care should serve, and how care might appropriately proceed in the particular situation. Through this dialogue, a shared understanding of the reconsidered clinical situation could emerge and provide grounds for subsequent clinical judgment, decision, or response.

Clinical responses involved determining how care should proceed in light of what had been reconsidered. This could take the form of action, non-action, a newly shared understanding, a reshaped clinical relationship, continuing care, or some combination of these. In some encounters, the ensuing course of care remained unchanged, while the grounds on which it was understood and judged appropriate for the particular patient had changed.

Provisional closure was reached when reconsideration had provided sufficient grounds for a clinical response, that response had been determined, and what had been reopened could therefore assume a provisionally settled status. Patient reality thus functioned not only in giving rise to the consequential mismatch that prompted reopening, but as an ongoing constraint on the adequacy of the understanding and response that followed. We describe this recurrent process as Clinical Epistemic Reopening (CER): consequential mismatch → inquiry into medical framing → reopening → reconsideration → clinical response → provisional closure

These components were analytically distinguishable, although in practice they could overlap and unfold within the same clinical dialogue. The application of this analytic framework across the 31 cases is summarized in Supplementary Table S2.

## 4. Discussion

### 4.1. Clinical Epistemic Reopening

This study identified Clinical Epistemic Reopening (CER), a recurrent clinical process through which medical framing can itself become reconsiderable when its adequacy for a particular patient is called into question by that patient’s reality. Medical science makes reality clinically knowable by reducing it; CER makes the resulting framing selectively reconsiderable when patient reality calls its adequacy into question. CER therefore neither opposes scientific reduction nor simply supplements it with contextual or experiential information from outside. Rather, it allows the framing produced through reduction itself to become an object of clinical inquiry.

Dialogue supported this process throughout. As patients explored the concerns they brought to the consultation, dialogue helped clarify whether their concerns could be addressed within existing medical framing or whether a consequential mismatch called its adequacy into question, prompting inquiry into what within that framing required reopening. Dialogue then enabled the clinician and patient to work with what had become reconsiderable in relation to the patient’s reality, contributing to the grounds on which a clinical response could be determined and provisional closure reached.

The issue arises not primarily at the level of whether generalized medical knowledge is valid, but at the point where that knowledge is brought to bear on the reality of a particular patient. Medical knowledge may retain its warrant while aspects of medical framing become reconsiderable, including how the clinical problem is framed, what medical knowledge can legitimately determine, how its authority should be understood, and what purposes it should serve in relation to a particular patient. CER does not prescribe the direction of the ensuing response; rather, it changes what can become relevant to the grounds on which that response is clinically judged.

CER thus extends clinical reasoning beyond determining what should be done within an established medical framing: when warranted by patient reality, the adequacy of that framing can itself become an object of clinical inquiry. In this sense, CER can reverse the usual direction of clinical scrutiny: medical framing, which ordinarily functions as means of understanding and acting upon the patient, may itself come under scrutiny when patient reality calls its adequacy in the particular situation into question. This does not require medicine to abandon the reduction and provisional closure on which clinical practice depends; it allows the relevant aspect of an otherwise warranted framing to be selectively reopened when its adequacy becomes uncertain for a particular patient. The necessary selectivity of scientific medicine need not define the limits of its clinical reach.

### 4.2. Interdisciplinary resources in Clinical Epistemic Reopening

A broad interdisciplinary body of scholarship has examined medicine from different vantage points. Medical anthropology, sociology, philosophy, ethics, nursing scholarship, and the medical humanities have explored how medicine understands human phenomena, how medical knowledge and categories are constituted and applied, and what particular ways of framing illness and clinical problems make visible or leave less visible. Across their substantial differences, these traditions provide intellectual resources that may become relevant to the process of CER (Kleinman, 1988; Good, 1994; Rosenberg, 2002; Carel and Kidd, 2014; Schwab, 2012).

More specifically, in CER, interdisciplinary perspectives may serve not only to broaden understanding of the patient, but also to make medicine itself more intelligible within the clinical encounter-its assumptions, historical contingency, epistemic limits, purposes, and consequences. Such perspectives can illuminate what has become salient in the encounter, including dimensions of meaning, experience, values, social worlds, embodiment, temporality, or the limits of knowledge.

CER may therefore provide a clinical point of encounter at which scientific medicine and diverse forms of human inquiry can be brought into relation with one another. What biomedicine can reliably establish can retain its warrant while being brought into relation with forms of understanding that address questions biomedicine does not by itself settle. Which forms of knowledge become relevant depends on what has become clinically salient in the particular encounter. Interdisciplinary knowledge can thereby become clinically relevant where it helps clarify what medical knowledge can establish, what it means in a particular situation, where its authority reaches, and how it should be brought to bear in the life of a particular person.

### 4.3. What CER makes possible

Conceptualizing CER widens what can become available to clinical reasoning without abandoning medically warranted knowledge; the process remains directed toward determining a clinical response and reaching provisional closure.

Importantly, CER remains anchored in clinical practice. It begins not from an abstract critique of medical framing, but from a consequential mismatch arising in the encounter with a particular patient. CER thus allows clinical inquiry to extend into epistemological, ethical, or other forms of human inquiry without becoming detached from the practical demands of caring for the patient before the clinician.

For patients, CER may offer medicine a way of responding to patient dissatisfaction arising from concerns that remain unresolved despite medically appropriate care, by bringing those concerns into clinical reasoning rather than leaving them at its margins. The resulting clinical response may or may not involve a change in treatment or decision; its significance may also lie in how the course of care is understood, adjusted, or continued.

For clinicians, CER may provide a way of making sense of encounters in which medically warranted reasoning remains appropriate within its own terms yet seems insufficient in relation to the person before them. CER provides a conceptual language through which the disquiet arising in such encounters can become a reason for inquiry rather than remaining only an individual source of concern. What might otherwise be experienced as compassion, discomfort, or moral concern may also carry epistemic significance when it points to a consequential mismatch. Experienced or compassionate clinicians may sometimes arrive intuitively at responses similar to those described here. CER makes explicit the clinical reasoning through which such responses can arise when patient reality calls the adequacy of medical framing into question, allowing that reasoning to become examinable, discussable, and potentially teachable. CER may also allow medicine to learn cumulatively from consequential mismatches. Recurrent mismatches across encounters could help identify points that may warrant particular attention in routine clinical practice, expanding clinicians’ capacity to recognize where established medical framing may not fully account for a patient’s situation. Whether a mismatch is present and what, if anything, requires reopening would remain to be determined in relation to the particular patient.

### 4.4. Implications for clinical practice

CER does not imply that medical framing should routinely be reopened. The practical challenge is not to maintain continuous openness, but to preserve the capacity to notice when a consequential mismatch may be present and, when it is, to inquire into what requires reopening. The Dialogue-Based Outpatient Clinic may have facilitated such noticing by creating a space within medicine in which patients could bring concerns without a predefined agenda and what emerged in relation to those concerns could remain clinically relevant without being presumed to conform to an existing medical framing. In routine encounters, the attention of both patients and clinicians is necessarily directed toward understanding, explaining, deciding upon, and carrying out medically defined processes. This orientation is indispensable to clinical care, but may leave limited room for either party to move beyond the medical framing organizing the encounter and consider questions for which other forms of inquiry may become relevant. Separating the Dialogue-Based Outpatient Clinic from routine care may have created such room within medical care, including room to reflect on medicine itself. The interval before the scheduled consultation may also have given patients time to consider what they wished to discuss.

Within this space, clinicians could listen closely to patients’ concerns, explore them in greater depth, and remain with what emerged rather than moving quickly toward a predefined clinical task or response. This depth of engagement may have allowed consequential mismatches to become more fully apparent and provided room for clinicians and patients to pursue the inquiry and reconsideration that followed. It may also have allowed perspectives beyond biomedicine to become clinically relevant where they helped illuminate what had emerged in the encounter. A dedicated clinic, however, is neither constitutive of CER nor established by this study as necessary for it. What may matter is that somewhere within care there remains a possibility for patients to bring forward concerns when something no longer fits, and for clinicians to attend closely enough for a consequential mismatch to become recognizable. Patient-centered, narrative, and shared decision-making approaches may support the engagement with patient reality through which such mismatches become recognizable. Such approaches do not necessarily entail reopening medical framing, but they may provide conditions in which its adequacy can come into question and CER can arise.

At present, it remains unclear how readily the conditions that enabled CER in this setting can be reproduced within routine clinical care, or how reopening might be incorporated into such care while preserving the settledness required for clinical judgment and action. At least until greater clinical understanding and experience of CER have accumulated, the dedicated clinical space provided by the Dialogue-Based Outpatient Clinic may therefore offer a clinically workable way of allowing reopening where it is warranted while preserving the settledness on which routine clinical care also depends.

### 4.5. Limitations

Several limitations should be considered. First, this study was conducted in a single Dialogue-Based Outpatient Clinic within a breast oncology service and involved a relatively small number of patients. CER was developed within this particular clinical setting, patient population, and healthcare context, and its transferability to other specialties, settings, clinicians, and healthcare systems requires further investigation. Patients attending this clinic were also a self-selected or clinically referred group who may have been more likely than patients in routine oncology care to bring concerns unresolved within existing medical framing. The proportion of patients in whom CER occurred should therefore not be interpreted as an estimate of its prevalence in breast oncology or clinical practice more broadly.

Second, the physician who established the clinic conducted the consultations and led the initial analysis. This provided access to the clinical reasoning and longitudinal context through which the encounters unfolded, but it also means that what became visible in the consultations and how it was subsequently interpreted may have been shaped by the same clinician. Repeated comparison across cases, examination of contrasting cases in which existing framing remained adequate, consideration of alternative interpretations, and critical discussion within the research team were used to strengthen reflexivity, but cannot remove this limitation.

Third, the data consisted of routine clinical documentation and supplementary records prepared during and after consultations rather than audio-recorded, verbatim transcripts. These materials captured clinically consequential concerns, reasoning, and responses across encounters, but did not permit fine-grained analysis of the moment-to-moment interactional processes through which consequential mismatches became visible or inquiry, reopening, and reconsideration unfolded in dialogue.

Fourth, this was a theory-generating study based on clinical encounters in a particular setting. Although CER was identified as a recurrent process across these encounters, how it may be recognized and understood by other clinicians in different clinical contexts remains to be explored.

Finally, this study did not seek to establish a fixed taxonomy of the forms of medical framing implicated in consequential mismatches. Further comparative analysis may clarify recurrent patterns and relationships among them, but any such analytic distinctions would themselves necessarily be selective and should not be treated as exhaustive categories into which future encounters must fit. What requires reopening would still need to emerge through engagement with the particular patient in the particular clinical encounter.

## 5. Conclusion

Scientific medicine gains much of its power by reducing complex human realities into forms that make knowledge, judgment, and action possible. Such reduction is necessarily selective, and the framing it produces may remain medically warranted even when its adequacy in relation to the reality of a particular patient can no longer simply be assumed. CER describes a clinical process through which the relevant medical framing can then become reconsiderable in relation to that patient’s reality.

This does not require medically warranted knowledge to be weakened or medical framing to remain perpetually open. Rather, it requires preserving the possibility of recognizing when patient reality calls the adequacy of existing medical framing into question, inquiring into what requires reopening, and reconsidering what has been reopened in relation to that patient. CER therefore allows clinical inquiry, when warranted by patient reality, to turn toward medicine itself rather than remaining directed only toward the patient.

CER therefore locates human reality neither outside scientific medicine nor in opposition to it. Patient reality can become relevant to the adequacy of the framing through which medical knowledge is brought to bear, while what that knowledge can legitimately establish retains its warrant. CER thus allows the capacities of scientific medicine to remain fully available while making the framing through which they are applied selectively reconsiderable in relation to patient reality. The necessary selectivity of scientific medicine need not define the limits of its clinical reach.

**Figure 1.**
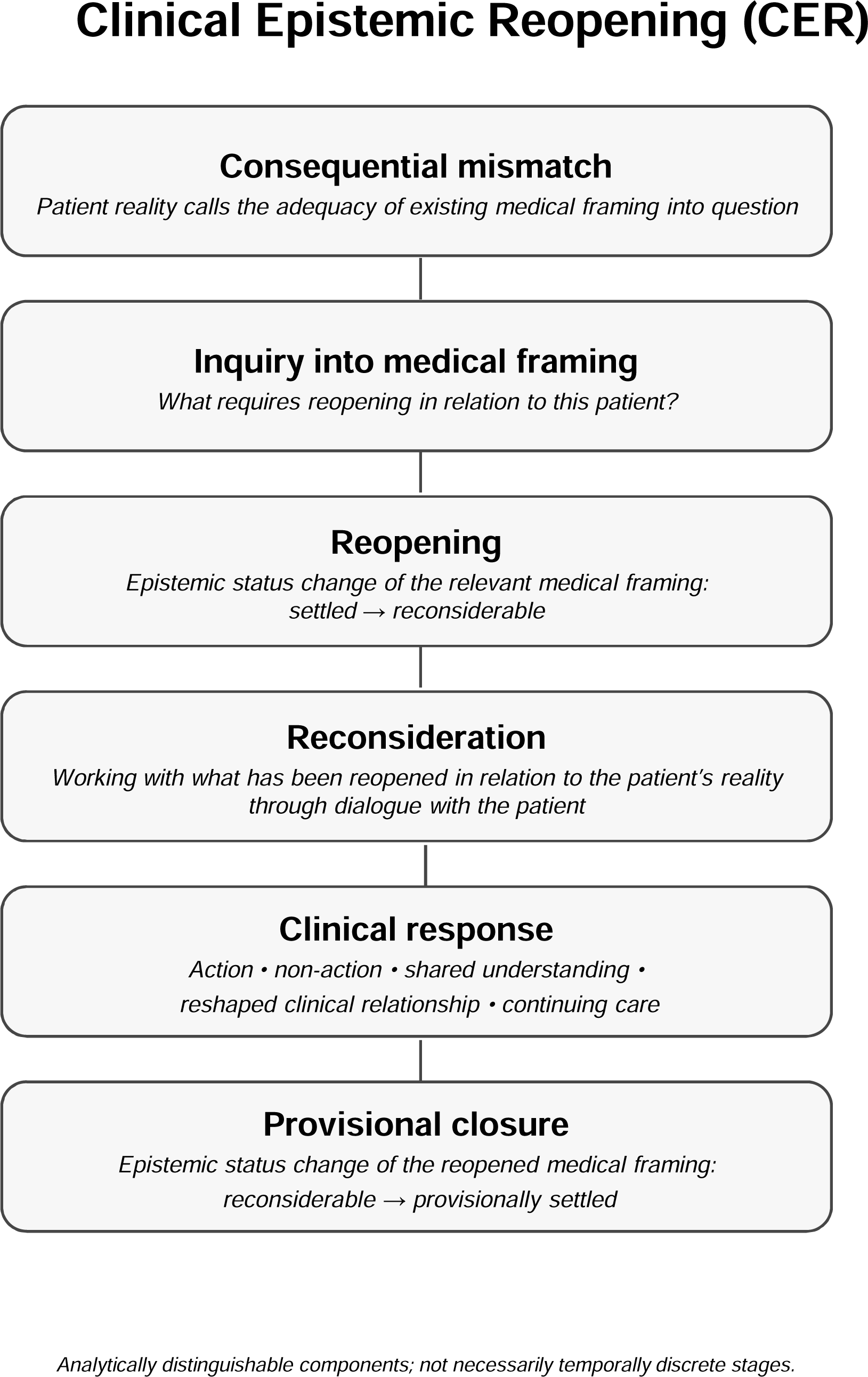
Clinical Epistemic Reopening (CER). A consequential mismatch becomes apparent when patient reality calls the adequacy of existing medical framing into question. The mismatch prompts inquiry into medical framing to determine what requires reopening in relation to the particular patient. This inquiry may extend from the framing in which the mismatch first appeared to related or more fundamental framing. Reopening changes the epistemic status of the relevant medical framing from settled to reconsiderable. Reconsideration involves working with what has been reopened in relation to the patient’s reality through dialogue with the patient. Clinical response involves determining how care should proceed in light of what has been reconsidered and may take the form of action, non-action, a newly shared understanding, a reshaped clinical relationship, continuing care, or some combination of these. Provisional closure is reached when reconsideration has provided sufficient grounds for a clinical response, that response has been determined, and the reopened medical framing can assume a provisionally settled status. These components are analytically distinguishable but need not unfold as temporally discrete stages. Provisional closure remains subject to the patient’s evolving reality; a subsequent consequential mismatch may initiate further inquiry and reopening.

## Supporting information

Supplement aryTable2

## Data Availability

The data underlying this study consist of clinical records containing confidential patient information and are not publicly available due to ethical and privacy restrictions. De-identified case-level summaries supporting the analysis are provided in the Supplementary Tables.

