## Supplementary Table1 for "Clinical Epistemic Reopening: When Patient Reality Makes Medical Framing Reconsiderable"

**Supplementary Table S1**

**Cases in which existing medical framing remained adequate**

**Purpose of the table**

**This table presents the 13 patients whose concerns could be addressed through biomedical information, clarification, investigation, referral, or other clinical responses while the medical framing remained adequate. These cases were retained in the analysis as contrasts to encounters in which a consequential mismatch made some aspect of existing medical framing reconsiderable.**

| **Case** | **Concern brought to the clinic** | **Clinical response** |
| --- | --- | --- |
| **1** | **The patient requested detailed information about breast reconstruction surgery.** | **The surgical procedure, expected postoperative course, and potential complications were explained.** |
| **2** | **Following a diagnosis of breast cancer, the patient asked about the standard postoperative treatment for her type of breast cancer.** | **The patient was informed that radiotherapy, chemotherapy, and endocrine therapy would be considered according to the postoperative findings, and the approximate duration of treatment was explained.** |
| **3** | **Following a diagnosis of breast cancer, the patient asked about the expected course of standard treatment.** | **The patient was informed that, depending on the postoperative findings, Oncotype DX testing could be considered if desired, and that radiotherapy, chemotherapy, and endocrine therapy might subsequently be considered. The approximate duration of treatment was also explained.** |
| **4** | **Following a diagnosis of breast cancer, the patient asked about the expected course of standard treatment and whether isoflavone intake was advisable.** | **The expected postoperative treatment pathway, including possible radiotherapy, chemotherapy, and endocrine therapy according to the postoperative findings, and the approximate duration of treatment were explained. Information concerning isoflavones contained in relevant clinical guidelines was also provided.** |
| **5** | **While receiving adjuvant endocrine therapy after breast cancer surgery, the patient asked about the guideline-recommended duration of treatment.** | **The recommendations concerning the duration of endocrine therapy were explained.** |
| **6** | **During follow-up after breast cancer surgery, the patient reported occasional headaches. An otolaryngological assessment had not identified sinusitis, and she asked which specialty she should consult next. Further evaluation was needed, including consideration of possible brain metastasis.** | **Consultation with a neurologist was recommended for further evaluation.** |
| **7** | **While receiving endocrine therapy after breast cancer surgery, the patient was concerned about possible loss of bone density and wished to have it assessed.** | **Bone mineral density was measured and confirmed to be within the normal range.** |
| **8** | **Following a diagnosis of breast cancer, the patient reported a persistent cough and asked about its cause.** | **Diagnostic testing identified COVID-19 infection as the cause of the symptom, and appropriate treatment was initiated.** |
| **9** | **Several years after breast cancer surgery, the patient wished to know the subtype of her breast cancer.** | **The tumor subtype was confirmed from the medical record and explained to the patient.** |
| **10** | **Following breast cancer surgery, the patient had been advised by her treating physician to take oral anticancer therapy and asked whether available evidence actually showed a benefit from taking the medication.** | **The evidence indicating improved outcomes with the treatment was explained. After receiving this information, the patient decided to take the medication.** |
| **11** | **Following a diagnosis of breast cancer, the patient sought more detailed information about how her planned breast-conserving surgery would be performed than had been provided by her treating physician.** | **The planned breast-conserving procedure was explained, using ultrasonography to show the location of the tumor and approximate surgical markings.** |
| **12** | **Following a diagnosis of breast cancer, the patient asked general questions about postoperative daily life, the cost of surgery, and subsequent treatment.** | **An overview of postoperative life, surgical costs, and the anticipated course of subsequent treatment was provided.** |
| **13** | **Following breast cancer surgery, the patient reported calf pain.** | **Ultrasonography was performed to assess for possible deep-vein thrombosis. No evidence of deep-vein thrombosis was identified, and the result was explained to the patient.** |
