## Supplement aryTable2 for "Clinical Epistemic Reopening: When Patient Reality Makes Medical Framing Reconsiderable"

**Supplementary Table S2**

*Case-level analytic mapping of Clinical Epistemic Reopening (CER)*

This table presents a case-level analytic mapping of the clinical process of CER across the 31 cases in which consequential mismatches were identified. To protect patient confidentiality, case descriptions have been deliberately limited to information necessary to show the clinical and epistemic structure relevant to CER. Potentially identifying demographic, clinical, and contextual details have been omitted or generalized. Accordingly, the cases do not provide comprehensive clinical histories or exhaustive accounts of the encounters. In some cases, this necessary reduction of contextual detail may limit the reader’s ability to fully reconstruct why a particular aspect of medical framing became the object of reopening. The table therefore presents the resulting structure of CER rather than a complete reconstruction of the clinical reasoning through which it emerged.

**Case 1**

| **Analytic component** | **Concise case-level account** |
| --- | --- |
| **Patient reality / consequential mismatch** | A patient did not want the diagnosis of cancer explicitly named because it had become associated, through a previous experience of serious illness in someone close to her, with catastrophic decline and loss of independence. The clinical pathway depended on diagnosis while ordinarily assuming that the diagnosis would be explicitly communicated. |
| **Existing medical framing** | Clinical care ordinarily proceeds by identifying a disease through diagnosis, using that diagnosis to guide clinical reasoning and treatment decisions, communicating that diagnosis to the patient, and organizing treatment and subsequent care accordingly. |
| **Reopening / reconsideration** | Medicine’s own way of reasoning, and the relationship between that reasoning and how its knowledge and treatments are made available to patients, became objects of inquiry. This inquiry rendered the assumed linkage between diagnosis, its disclosure to the patient, and the provision of treatment reconsiderable. Reopening these assumptions allowed medicine’s use of diagnostic categories to represent disease and organize clinical reasoning and treatment to be distinguished from whether the patient herself had to take up the diagnostic label, and the meanings it carried for her, as the terms in which she understood her condition in order to receive medically warranted treatment. The clinician asked whether she wished to receive medically warranted treatment without adopting medicine’s diagnostic framing as her own. |
| **Clinical response / provisional closure** | She chose to proceed with treatment, understanding her condition not through the diagnostic label, but more concretely as a growing mass in her breast. The reopened relationship between medicine’s diagnostic representation and its clinical reasoning and treatment was provisionally closed on this basis. |

**Case 2**

| **Analytic component** | **Concise case-level account** |
| --- | --- |
| **Patient reality / consequential mismatch** | No residual cancer was found after mastectomy following neoadjuvant chemotherapy, representing oncological success within the existing medical framework. However, a gap emerged between a treatment course that remained medically warranted and the patient’s experience of irreversible breast loss when no cancer was ultimately found. This gap was intensified both by the characterization of treatment as successful and by the fact that medical professionals approached her continuing suffering primarily in psychiatric terms. |
| **Existing medical framing** | Pathological complete response represents a highly favorable oncological outcome. Persistent distress after successful cancer treatment may subsequently be framed primarily as a psychological or psychiatric problem. |
| **Reopening / reconsideration** | Medicine’s own way of reasoning and its representation of treatment success became objects of inquiry. This inquiry extended to what medicine is and how medicine has been formed historically. The framing reopened was that, while medical knowledge and reasoning ordinarily function as a warranted basis for clinical judgement and treatment, modern medical practice does not ordinarily provide a way for clinicians and patients to share an understanding of medicine itself – its reasoning, possibilities, limitations, historical situatedness, and the basic concepts that underlie it. The clinician shared with the patient why the treatment had been considered appropriate at that time and, by tracing the history of breast surgery, how medical treatment had been historically formed, with its changing possibilities and limitations. Their discussion extended to what medicine itself is. |
| **Clinical response / provisional closure** | Through this discussion, she came to understand that the healthcare professionals who had treated her had also been working for her within the medical framework available to them. She said that, while hoping that future medicine would not remain as it was, she could now move forward. She situated her loss within the medical reasoning and limitations surrounding her treatment, and breast reconstruction became a meaningful possibility. |

**Case 3**

| **Analytic component** | **Concise case-level account** |
| --- | --- |
| **Patient reality / consequential mismatch** | Repeated experiences had shown the patient that what she perceived in her body could lose clinical weight when it was not captured by established tests or measurements. Through these experiences, she had become aware that what medicine makes available as clinical knowledge may represent only selected aspects of bodily reality. This became consequential when she later faced a decision about whether to undergo surgery for a head and neck tumor whose benign or malignant nature could not be determined. The discrepancy made examinable not only particular findings but the selective way in which medicine makes bodily reality available for clinical knowledge. |
| **Existing medical framing** | Clinical medicine makes bodily phenomena knowable and actionable by selecting particular aspects of reality for observation, measurement, classification, and analysis. Symptoms are investigated through clinically defined questions and established methods, and the resulting findings provide warranted grounds for clinical judgment and action. |
| **Reopening / reconsideration** | Medicine’s way of knowing by selecting aspects of reality for observation, measurement, and analysis—and the reach of the understanding produced through that selection—became reconsiderable. The patient’s previous experiences were reconsidered as instances in which medically warranted forms of observation had captured particular aspects of her condition without necessarily exhausting what could be clinically relevant. This shared understanding allowed her bodily perceptions to retain clinical significance without displacing established medical assessment, providing a basis for future concerns to be examined through both what medicine could establish and what might remain insufficiently captured by its existing frame. |
| **Clinical response / provisional closure** | With this shared understanding of what medicine could and could not establish, the patient and clinician considered whether she should undergo surgery for the head and neck tumor, which carried a risk of complications. Recognizing medicine’s limitations and incompleteness, she chose to trust what medicine had come to know through its historical development and decided to undergo surgery. She subsequently experienced facial nerve palsy, a postoperative complication, which gradually resolved. |

**Case 4**

| **Analytic component** | **Concise case-level account** |
| --- | --- |
| **Patient reality / consequential mismatch** | A patient after breast cancer surgery questioned whether she should accept the burdens of radiotherapy and endocrine therapy on her present body and life in order to reduce the risk of a recurrence that had not yet occurred. The mismatch arose between medicine’s orientation toward preventing future disease through interventions that impose burdens on the body in the present and the patient’s concern about what that preventive aim could justify doing to her body now. |
| **Existing medical framing** | Adjuvant therapies are recommended on the basis of medical evidence to reduce the future risk of recurrence. Within this framing, intervention in the present is medically justified by its expected reduction of future disease risk. |
| **Reopening / reconsideration** | The framing that prevention of future disease can justify medical intervention in the present body became reconsiderable. The clinician and patient shared an understanding that medicine can estimate future risks and benefits, but does not itself provide a measure of the value of the present moment in the patient’s life. The medical rationale for reducing future recurrence remained valid, while its authority to justify intervention in the patient’s present body was no longer treated as self-sufficient. Adjuvant treatment was then reconsidered in relation to both the future it sought to protect and the present on which it would impose its burdens. |
| **Clinical response / provisional closure** | After reconsidering adjuvant treatment in relation to both the future it sought to protect and the present on which it would impose its burdens, she chose to undergo treatment. |

**Case 5**

| **Analytic component** | **Concise case-level account** |
| --- | --- |
| **Patient reality / consequential mismatch** | A patient choosing between two medically reasonable surgical approaches repeatedly sought confirmation that one was the medically “right” decision. Both surgical options were medically reasonable, yet the patient sought a medically “right” choice that medical knowledge could not determine because it could not predict which individual future would follow. |
| **Existing medical framing** | When more than one medically reasonable option is available, comparative evidence, expected benefits and harms, and patient preferences are ordinarily used to support an individual treatment decision. |
| **Reopening / reconsideration** | The aspect of medical framing that became reconsiderable was what could ground an individual decision when medical knowledge itself could not determine the choice: what criteria of value, not encompassed by medical determination, could legitimately provide a basis for deciding between medically warranted options. The clinician and patient first made explicit what medical knowledge could and could not determine between the two medically warranted options. They then constructed an explicit account of what was not encompassed by biomedical determination and considered what values within that domain could legitimately ground a clinical decision. |
| **Clinical response / provisional closure** | Once the limits of medical determination had been made explicit, the patient identified avoiding future regret as the value judgment that would ground the decision medicine itself could not determine and, on that basis, chose bilateral mastectomy. |

**Case 6**

| **Analytic component** | **Concise case-level account** |
| --- | --- |
| **Patient reality / consequential mismatch** | A patient recommended neoadjuvant chemotherapy believed that her breast cancer might have resulted from insufficient attention to diet and daily rhythms and wished henceforth to put only what she regarded as good into her body. She therefore found it difficult to accept introducing cytotoxic treatment into her body and expressed a broader reluctance to rely on science. The mismatch was not resolved by explaining the expected benefits and adverse effects of chemotherapy, because her concern was directed at the form of medicine itself: treating disease by acting upon the body through interventions that may themselves be harmful. |
| **Existing medical framing** | Scientific medicine identifies disease processes through forms of knowledge that make them available for investigation and intervention, and evaluates treatments according to their capacity to alter those processes and improve clinically relevant outcomes. Within this framing, treatment-related harm may be warranted when it is proportionate to the expected therapeutic benefit. |
| **Reopening / reconsideration** | The framing that became reconsiderable was scientific medicine’s underlying relation to disease and the body: it produces knowledge by analytically isolating disease processes and seeks to alter them through interventions on the body, even when those interventions may themselves cause harm. The clinician made this form of medicine explicit and acknowledged the patient’s concern about it rather than treating scientific medicine as a self-evident authority. They then reconsidered what this form of medicine could offer in her particular situation. If cure of the breast cancer was taken as the primary aim, scientific medicine provided the most warranted basis then available for pursuing that aim, making chemotherapy a meaningful option despite her continuing reservations about its mode of intervention. |
| **Clinical response / provisional closure** | The patient did not need to abandon her concern about scientific medicine in order to accept treatment. Having reconsidered what scientific medicine could offer in relation to the aim of cure, she chose to undergo chemotherapy. |

**Case 7**

| **Analytic component** | **Concise case-level account** |
| --- | --- |
| **Patient reality / consequential mismatch** | A patient described how a postoperative complication involving her surgical wound had created a rupture in her marital relationship that had never been repaired and was now approaching its breakdown. The surgical wound had resolved as a medical problem, while the patient experienced consequences that she understood as having extended from the intervention into her sexuality, marital relationship, and subsequent life. She also felt that she had been unable to speak about such consequences within her medical care, including with her treating physician and other healthcare professionals. |
| **Existing medical framing** | Breast cancer surgery is ordinarily evaluated in terms of oncological adequacy, procedural outcomes, complications, recovery, and recognized treatment-related morbidity. Once a surgical wound and its recognized physical effects have resolved medically, consequences extending into the patient’s subsequent relational and biographical life may fall outside the clinical problem organized around cancer treatment. |
| **Reopening / reconsideration** | The aspect of medical framing that became reconsiderable was the scope of medicine’s responsibility for the consequences of its own interventions: what responsibility medicine retained after medically recognized complications had resolved. The clinician and patient considered how an invasive medical intervention can have consequences that extend beyond medically recognized complications into sexuality, intimate relationships, and the subsequent course of a person’s life. They also considered what medicine could and could not take responsibility for within such wider consequences, without reducing them either to a surgical complication or to matters entirely outside clinical concern. |
| **Clinical response / provisional closure** | The physician acknowledged that the effects of medical intervention can extend beyond what medicine ordinarily represents as treatment-related morbidity and recognized the patient’s account as clinically meaningful. The patient said that, in this clinic, she had been able to bring into clinical dialogue experiences that she had previously been unable to speak about elsewhere within her medical care. |

**Case 8**

| **Analytic component** | **Concise case-level account** |
| --- | --- |
| **Patient reality / consequential mismatch** | A patient faced a treatment recommendation that was medically warranted and supported within national health and the hospital system, but did not feel that the recommended course corresponded to how she understood her situation or wished to proceed. The treatment remained medically warranted, yet the expectation that she should follow the recommended course became consequential when her own understanding and intended course diverged from it. The patient experienced the recommendation as reinforced not only by clinical reasoning but also by the institutional system surrounding her care, contributing to a sense that she was expected to follow the recommended course. |
| **Existing medical framing** | National health and healthcare institutions organize care around medically warranted knowledge, standards, and treatment pathways. Within hospitals, treatments supported by established medical knowledge are translated into recommendations and systems of care that facilitate their implementation. |
| **Reopening / reconsideration** | The aspect of medical framing that became reconsiderable was the institutional authority through which medically warranted treatment is organized and reinforced within the healthcare system, and the extent to which that authority could legitimately determine the course of treatment for an individual patient. The clinician and patient distinguished the medical warrant for the recommended treatment from the additional authority it acquired through its institutional implementation. The treatment could remain medically justified and appropriately supported by the healthcare system without thereby establishing that the patient was required to follow it. |
| **Clinical response / provisional closure** | The institutional authority supporting the recommended treatment was distinguished from an obligation for her to follow that course. With this understanding of the choice available to her, she chose for the time being to proceed with the guideline-based recommendation. |

**Case 9**

| **Analytic component** | **Concise case-level account** |
| --- | --- |
| **Patient reality / consequential mismatch** | A patient described how the diagnosis of breast cancer had come to signify more than the medically defined disease itself. The diagnostic category remained medically valid, but the meaning carried by the words “breast cancer” in the patient’s reality extended beyond what the diagnosis itself medically established. A term that functioned within medicine as a clinically useful representation of disease had come to signify a broader and continuing threat, affecting how the patient and her family understood her body, future, and subsequent choices. |
| **Existing medical framing** | Diagnostic language enables medicine to identify, classify, and communicate disease in clinically meaningful categories. A diagnosis such as breast cancer brings complex biological findings together under a shared medical designation through which prognosis, treatment, surveillance, and subsequent clinical decisions can be organized. |
| **Reopening / reconsideration** | The aspect of medical framing that became reconsiderable was the representational function of diagnostic language: medically meaningful categories can come to represent more of a patient’s life than they medically establish. The clinician and patient distinguished the medical meaning of the diagnosis from the broader meanings that the diagnostic label had acquired in her life. Breast cancer remained a valid medical diagnosis, while what that diagnosis should mean for her identity, future, and subsequent choices was not determined by the diagnosis itself. |
| **Clinical response / provisional closure** | The patient and clinician developed a shared understanding of what the diagnosis could medically establish and what remained undetermined by the diagnostic label. This allowed subsequent decisions, including those concerning reconstruction, to be considered without treating the broader meanings attached to “breast cancer” as medically given. |

**Case 10**

| **Analytic component** | **Concise case-level account** |
| --- | --- |
| **Patient reality / consequential mismatch** | A patient was concerned about breast cancer recurrence and wished to undergo imaging surveillance. Her treating physician had considered such imaging unnecessary in accordance with guideline-based surveillance recommendations, and she sought consultation because she remained uncertain whether imaging might nevertheless have value for her. |
| **Existing medical framing** | Post-treatment surveillance is ordinarily guided by population-level evidence regarding whether routine investigations improve clinically meaningful outcomes. When routine imaging in asymptomatic patients has not been shown to improve prognosis, it is generally not recommended as part of standard surveillance. |
| **Reopening / reconsideration** | What became reconsiderable was whether the absence of demonstrated prognostic benefit was sufficient to conclude that surveillance imaging had no clinical value for an individual patient. The clinician and patient distinguished between what the evidence established and the further clinical judgment made from it. The absence of demonstrated prognostic benefit remained medically warranted knowledge without determining whether imaging could contribute to a better outcome for this individual patient. |
| **Clinical response / provisional closure** | The patient understood that routine imaging had not been shown to improve prognosis in asymptomatic patients and that CT could not be presented as a proven means of improving survival. At the same time, she understood that, in an individual patient, imaging might detect a lesion that could be treated and potentially contribute to survival, even though such a benefit had not been demonstrated at the population level. She wished to undergo imaging to determine whether there was a lesion in her body that would warrant medical intervention. |

**Case 11**

| **Analytic component** | **Concise case-level account** |
| --- | --- |
| **Patient reality / consequential mismatch** | A patient described how, during postoperative radiotherapy, her spouse had avoided physical proximity because he believed that radiation associated with her treatment could harm him. She experienced this as deeply painful. Although radiotherapy had been medically indicated and completed as intended, an intervention directed at treating her cancer had acquired meanings outside its biomedical purpose and had consequentially affected her closest relationship. This consequence was not represented among the recognized medical outcomes or adverse effects of treatment. |
| **Existing medical framing** | Postoperative radiotherapy is ordinarily represented through its oncological indication, expected reduction in recurrence risk, treatment delivery, and recognized adverse effects. Once treatment is completed, its clinical evaluation may close around these biomedical outcomes. |
| **Reopening / reconsideration** | What became reconsiderable was the extent to which the consequences of a medical intervention could be understood only through its intended biomedical effects and recognized adverse effects. Radiotherapy remained medically warranted, but the intervention had also acquired meanings in the patient’s relational world that medicine had neither intended nor ordinarily represented as an outcome of treatment. |
| **Clinical response / provisional closure** | The clinician acknowledged the patient’s suffering as a consequence connected to medical intervention. The patient’s account could thereby enter clinical dialogue as part of wider consequences through which a medical intervention had become consequential in her life, even though it was not a recognized adverse effect of radiotherapy. |

**Case 12**

| **Analytic component** | **Concise case-level account** |
| --- | --- |
| **Patient reality / consequential mismatch** | A patient with hormone receptor-positive breast cancer with lung metastasis had been told by her treating physician that cure was unlikely. Guideline-based treatment was therefore organized around long-term disease control and preservation of quality of life rather than cure. She asked whether this meant that cure was impossible for her as an individual. The medically warranted population-based framework supported treating metastatic breast cancer as generally incurable, but her question exposed a distinction between what could be warranted for a population and what could be known with certainty about an individual patient. |
| **Existing medical framing** | Metastatic breast cancer is generally regarded as treatable but not curable. For hormone receptor-positive disease without an immediate need for rapid tumor reduction, guideline-based treatment is ordinarily organized around long-term disease control, beginning with endocrine therapy. |
| **Reopening / reconsideration** | What became reconsiderable was the population-oriented logic through which medicine defines therapeutic aims, particularly the weight given to possibilities that occur only in a minority of patients. The clinician shared with the patient that, although metastatic breast cancer is generally considered incurable, prolonged complete remissions have been reported in a small minority of patients. They distinguished between what medicine could reasonably recommend on the basis of population-level outcomes and what those outcomes could establish about an individual patient. |
| **Clinical response / provisional closure** | The patient came to understand why standard treatment was organized around long-term disease control and why endocrine therapy was ordinarily recommended within that framework. With this understanding, she decided to discuss with her treating physician whether she might pursue a treatment strategy aimed at the possibility of cure rather than simply following the standard treatment pathway. |

**Case 13**

| **Analytic component** | **Concise case-level account** |
| --- | --- |
| **Patient reality / consequential mismatch** | A patient with multiple metastases, including gastric metastasis, asked whether her disease could be cured. She had been told that metastatic disease was generally considered incurable, but wanted to know whether this meant that cure was impossible for her as an individual. |
| **Existing medical framing** | Metastatic breast cancer is generally approached as treatable but not curable. Guideline-based treatment is therefore organized primarily around disease control, prolongation of survival, and preservation of quality of life rather than cure. |
| **Reopening / reconsideration** | What became reconsiderable was the population-oriented logic through which the low probability of cure becomes the basis for organizing standard therapeutic aims. The clinician and patient distinguished between the fact that cure is uncommon across populations and the stronger conclusion that cure was impossible for this individual patient. They also examined how this population-level expectation had shaped the therapeutic aims around which standard treatment was organized. |
| **Clinical response / provisional closure** | The patient came to understand why standard treatment was organized around disease control rather than cure, while also understanding that this population-level framework did not establish that cure was impossible for her individually. With this distinction made explicit, she decided to discuss with her treating physician what therapeutic aim she wished to pursue and whether treatment could be reconsidered in relation to that aim. |

**Case 14**

| **Analytic component** | **Concise case-level account** |
| --- | --- |
| **Patient reality / consequential mismatch** | A patient occasionally experienced pain at the surgical site, which brought back memories of her breast cancer and the treatment she had undergone. No new medical problem requiring intervention was apparent, yet the occasional pain brought the patient’s experience of breast cancer and treatment back into the present and led her to seek consultation in this clinic with a physician who had been involved in her previous cancer care and knew what she had been through. The relationship itself continued to matter to her despite the absence of a new biomedical problem to address. |
| **Existing medical framing** | Once breast cancer treatment has been completed and no serious pathology requiring further intervention is identified, the principal clinical episode may be considered closed. The physician–patient relationship is ordinarily organized around the diagnosis, treatment, and follow-up of clinically identifiable problems. |
| **Reopening / reconsideration** | What became reconsiderable was the role of the physician–patient relationship: whether it should be organized only around the diagnosis and treatment of medical problems, or whether a relationship formed through a shared treatment history could itself retain clinical meaning after the principal medical episode had ended. |
| **Clinical response / provisional closure** | No new medical intervention was required, but the continuing significance of the relationship with a physician who knew her treatment experience was recognized within clinical care. The consultation provided a place for that experience to be shared, and continued clinical availability was maintained should she wish to return. |

**Case 15**

| **Analytic component** | **Concise case-level account** |
| --- | --- |
| **Patient reality / consequential mismatch** | A patient had intermittent abdominal pain despite repeated medical evaluations at several hospitals without an identifiable cause. She had been told that nothing was wrong, yet the pain continued. Unable to reconcile the continuing pain with being told that there was no medical problem, she came to the clinic seeking guidance on how to understand and respond to it. |
| **Existing medical framing** | When repeated investigations do not identify a significant abnormality, diagnostic inquiry may close around the conclusion that no medical problem has been identified, and the clinical significance of continuing symptoms may remain unresolved. |
| **Reopening / reconsideration** | The reach of medical investigation itself became reconsiderable. The clinician and patient examined what negative investigations could and could not establish, recognizing that medical investigations examine particular aspects of clinical reality through specific methods and technologies rather than apprehending it in its entirety. Negative findings could therefore be understood within the limits of what those investigations could establish without negating the patient’s continuing pain. |
| **Clinical response / provisional closure** | What had been established, what remained unknown, and what could reasonably follow from that uncertainty were considered together with the patient. Continued observation became a reasonable way forward, with reassessment if symptoms changed or further concerns arose. |

**Case 16**

| **Analytic component** | **Concise case-level account** |
| --- | --- |
| **Patient reality / consequential mismatch** | A patient receiving neoadjuvant chemotherapy came to this clinic because, although continuation of chemotherapy was medically recommended, she was uncertain whether she had sufficient physical and mental strength to continue treatment and was struggling with whether she should proceed. |
| **Existing medical framing** | Her immediate clinical concern was whether she could continue chemotherapy. Information relevant to that question would ordinarily be organized around treatment toxicity, physical condition, and oncological concerns. |
| **Reopening / reconsideration** | Rather than taking the question whether chemotherapy could be continued as the fixed starting point of the consultation, the clinician listened to the patient’s concerns without restricting the discussion to what had already been defined as medically relevant. During an extended conversation in which she felt able to speak about what concerned her without first determining whether it was medically relevant, she gradually mentioned visual and shoulder symptoms that she had attributed to aging and therefore had not considered relevant to discuss with her treating physician. What became reconsiderable was medicine’s way of knowing the patient and the scope of what it could come to know through the clinical encounter. Experiences that had remained outside the existing medical framing thereby entered clinical reasoning and acquired medical significance. |
| **Clinical response / provisional closure** | The clinician explained that these symptoms could be medically significant and recommended further investigation. The patient agreed, and brain MRI was arranged. Brain MRI identified a cerebral infarction, for which she was admitted to the neurology service and treatment was initiated. |

**Case 17**

| **Analytic component** | **Concise case-level account** |
| --- | --- |
| **Patient reality / consequential mismatch** | A patient was found to have a small pulmonary lesion of uncertain nature and was advised to undergo surgery to establish a diagnosis. The purposes ordinarily organizing the proposed medical course did not fully coincide with what the patient wished her care to preserve. Establishing a diagnosis and treating possible malignancy could offer medical benefit, but pursuing these aims might also disrupt the independent life she valued and was currently able to maintain. |
| **Existing medical framing** | A pulmonary lesion suspicious for malignancy is ordinarily approached through diagnostic evaluation, including surgical diagnosis when indicated. Within this framing, establishing a diagnosis, treating disease where possible, and potentially prolonging life organize the proposed medical course. |
| **Reopening / reconsideration** | What became reconsiderable was the purpose organizing the proposed medical course: whether establishing a diagnosis, treating disease where possible, and potentially prolonging life should continue to organize care when these purposes did not fully coincide with the life the patient wished to preserve. The clinician and patient considered the uncertainties on both sides and shared that medicine could not determine in advance which course would ultimately prove least harmful. They therefore reconsidered not merely whether surgery should be performed, but what medicine was to be used for in her situation and, accordingly, which purposes should organize her care. |
| **Clinical response / provisional closure** | The patient chose surveillance, allowing the situation to be reconsidered over time if the lesion, her circumstances, or the priorities she wished medicine to serve changed. |

**Case 18**

| **Analytic component** | **Concise case-level account** |
| --- | --- |
| **Patient reality / consequential mismatch** | A patient with low-risk early breast cancer became particularly anxious about stopping endocrine therapy after a close friend died of breast cancer despite having received guideline-based treatment. Extended endocrine therapy was not routinely recommended in view of her low estimated risk of recurrence, while she understood the possible additional benefit to be uncertain rather than absent and placed particular value on reducing recurrence risk even if the expected benefit was small. |
| **Existing medical framing** | The duration of adjuvant endocrine therapy is ordinarily guided by evidence regarding recurrence reduction and treatment-related harms in defined patient populations. For patients at low risk of recurrence, the expected absolute benefit of extended therapy may be small, and extension beyond the standard duration is not routinely recommended. |
| **Reopening / reconsideration** | What became reconsiderable was not the validity of the statistical evidence underlying the recommendation, but the extent to which that evidence could determine the appropriate treatment duration for this individual patient. The clinician and patient examined the statistical basis of the recommendation, including recognized limitations of using population-derived treatment effects to inform decisions for an individual patient (Dahabreh, 2016). They also considered that guideline-based treatment represents the prevailing approach in contemporary medical practice, while medical interventions necessarily entail the possibility of harm, including adverse effects from extended endocrine therapy. Against this background, they examined what it meant for the patient to choose differently from the guideline-based recommendation, including understanding the uncertainty involved and taking responsibility for the choice and its possible consequences. |
| **Clinical response / provisional closure** | The patient understood both the statistical basis for the guideline-based recommendation and the uncertainty that remained for her individually, including the potential harms of extended therapy. With this understanding, she chose to continue endocrine therapy. |

**Case 19**

| **Analytic component** | **Concise case-level account** |
| --- | --- |
| **Patient reality / consequential mismatch** | A patient with severe persistent treatment-related peripheral neuropathy had substantial difficulty walking despite treatment from physicians in several specialties. Established therapeutic options had been exhausted, but the consequences of medical intervention had not ended for the patient. She continued to live in a body whose movement and everyday life had been substantially altered by treatment. |
| **Existing medical framing** | Treatment-related peripheral neuropathy is ordinarily assessed through symptoms, clinical findings, functional impairment, and response to established treatments. Once recommended therapeutic options have been exhausted without sufficient benefit, the condition may become framed as a persistent adverse effect for which medicine has little further treatment to offer. |
| **Reopening / reconsideration** | What became reconsiderable was whether the exhaustion of established therapeutic options could also mark the end of medicine’s responsibility, particularly when medical intervention itself had produced the patient’s continuing difficulty, or whether such harm required medicine to turn its attention back upon the consequences of its own intervention and remain clinically engaged with the reality it had helped create. The absence of an established treatment was distinguished from the absence of anything further for medicine to examine or respond to. |
| **Clinical response / provisional closure** | The patient and physician examined her gait together and experimented with cushioning and insole placement to reduce painful contact, resulting in visible improvement in gait and walking speed. Further low-risk approaches, including a topical preparation intended to improve local circulation, massage, and regular stretching and exercise, were considered with explicit recognition of their uncertain evidence and limitations. |

**Case 20**

| **Analytic component** | **Concise case-level account** |
| --- | --- |
| **Patient reality / consequential mismatch** | A patient with breast cancer had understood that, once a malignant lesion was identified and found to be surgically resectable, it should first be removed. She therefore found it difficult to understand why systemic therapy was recommended before surgery. The medically recommended sequence was therefore at odds with what she understood to be the more rational order of treatment. |
| **Existing medical framing** | The recommended treatment sequence was based on population-derived evidence. Administering systemic therapy before surgery also allows treatment response to be assessed and used to inform subsequent treatment decisions. On this basis, systemic therapy before surgery was medically recommended for patients meeting the relevant clinical criteria. |
| **Reopening / reconsideration** | What became reconsiderable was not whether the recommendation for neoadjuvant systemic therapy was medically warranted, but what its population-based rationale could establish about the preferable treatment sequence for this individual patient. The clinician and patient examined why systemic therapy before surgery was medically recommended, including how treatment response could inform subsequent treatment decisions, while making explicit both the population-based basis of that recommendation and the uncertainty that remained as to which sequence would ultimately prove better for this particular patient. |
| **Clinical response / provisional closure** | Although surgery first remained the more rational sequence from her own perspective, the patient chose to proceed with systemic therapy before surgery, accepting the medically recommended sequence while understanding both its population-based rationale and the uncertainty at the individual level. |

**Case 21**

| **Analytic component** | **Concise case-level account** |
| --- | --- |
| **Patient reality / consequential mismatch** | A patient in her 40s with advanced cancer no longer regarded prolongation of life or further disease-directed treatment as the primary aim of her care. Yet she wanted a physician who understood both her medical condition and how she wished to approach her life and death to remain involved as her disease progressed. Her concern therefore remained even after the question of further disease-directed treatment had largely been settled. |
| **Existing medical framing** | Biomedical care for advanced cancer is strongly organized around altering the course of disease and, where possible, prolonging survival. When these aims are no longer pursued, medical involvement may correspondingly be understood as receding or as being reorganized primarily around symptom control and end-of-life care. |
| **Reopening / reconsideration** | What became reconsiderable was the purpose of continued medical involvement and, with it, the physician’s role. The clinician and patient considered whether medical knowledge and the physician’s involvement had clinical meaning only insofar as they served disease-directed intervention, or what purposes continued medical involvement could serve when disease-directed intervention was no longer the primary aim. |
| **Clinical response / provisional closure** | The physician agreed to continue seeing her. Medical knowledge and clinical presence were thereby organized not primarily toward altering the disease course, but toward remaining medically involved as her condition progressed while attending to both her condition and the way she had chosen to approach her life and death. |

**Case 22**

| **Analytic component** | **Concise case-level account** |
| --- | --- |
| **Patient reality / consequential mismatch** | After completing long-term oncological follow-up, the patient returned because she wanted to speak once more with the physician who had cared for her over many years and to talk together about the time they had shared through her illness. No further oncological intervention or surveillance was required, yet she deliberately sought this final encounter with the physician who had accompanied her through that long course of care. |
| **Existing medical framing** | Within biomedical care, the physician’s role is principally organized around functions such as diagnosis, treatment, surveillance, and management of clinically defined problems. A long-term oncological relationship can therefore acquire a medically intelligible endpoint when these functions have been completed and further specialist follow-up is no longer indicated. |
| **Reopening / reconsideration** | What became reconsiderable was whether the physician-patient relationship was constituted only by the medical functions it had served. The patient’s return after those functions had reached their clinical endpoint brought into view that the relationship could retain clinical meaning beyond the medical functions through which it had been formed. |
| **Clinical response / provisional closure** | The encounter became a time for the patient and physician to acknowledge and reflect on the experience they had shared over ten years of illness and care. |

**Case 23**

| **Analytic component** | **Concise case-level account** |
| --- | --- |
| **Patient reality / consequential mismatch** | Having previously experienced serious illnesses, the patient sought a clinical encounter while she was well because she wanted a physician to know who she was in preparation for possible future illness. She anticipated that, if she became ill again, what a future symptom, change, or illness meant for her might not be fully understood from information gathered only after a medical problem had arisen. |
| **Existing medical framing** | Clinical encounters are ordinarily organized around a current medical problem, with information gathered according to its relevance to diagnosis, treatment, surveillance, or management. What is clinically important to know about a patient is therefore largely determined by the medical problem already requiring attention. |
| **Reopening / reconsideration** | What became reconsiderable was the temporal framing of clinical care as organized around medical problems once they have arisen. The patient’s concern opened the possibility that clinical involvement could begin before a new medical problem emerged, allowing the physician to know her while she was well so that this understanding could later inform care if illness or change occurred. |
| **Clinical response / provisional closure** | The physician came to understand why the patient wanted to be known while she was well and agreed to remain involved, allowing knowledge of her before future illness to become available if her condition later changed. |

**Case 24**

| **Analytic component** | **Concise case-level account** |
| --- | --- |
| **Patient reality / consequential mismatch** | After completing chemotherapy recommended by her physician, the patient questioned whether undergoing it had truly been a good choice for her. She had followed the medically recommended course, but recognized that completing a treatment regarded as appropriate did not allow her to know whether it had in fact been the better choice for her as an individual. |
| **Existing medical framing** | Cancer treatment is organized through disease categories and population-derived evidence that identify treatments expected to reduce recurrence risk. Once a diagnosis places a patient within a defined clinical category, guideline-based recommendations provide a medically warranted course of treatment on the basis of expected benefits and harms across comparable patients. |
| **Reopening / reconsideration** | What became reconsiderable was what it meant for a treatment decision to have been “right” when its justification was necessarily based on probabilistic knowledge. The clinician and patient distinguished between whether chemotherapy had been medically warranted at the time and whether anyone could know retrospectively that undergoing it had produced the better outcome for this particular patient. This distinction allowed the medical warrant for chemotherapy to be retained while bringing into clinical consideration a broader question that biomedical evaluation could not determine: what undergoing the treatment had meant within the patient’s life. |
| **Clinical response / provisional closure** | The clinician affirmed that the patient had completed a medically warranted treatment while acknowledging that whether it had produced the better outcome for her individually could not be known. They then considered the meaning of completing that treatment and how it might be valued within dimensions of her life not ordinarily captured by scientific medicine, including as something in which she could take pride. |

**Case 25**

| **Analytic component** | **Concise case-level account** |
| --- | --- |
| **Patient reality / consequential mismatch** | A patient with metastatic breast cancer wanted to be rechallenged with a therapy from which she had previously experienced substantial benefit, while her treating physician recommended moving to a new therapy. She sought consultation because she was uncertain which course to follow. |
| **Existing medical framing** | When metastatic breast cancer progresses during a systemic therapy, treatment is ordinarily changed to another therapy rather than returning to the treatment on which the disease had progressed. Subsequent treatment is selected from available options according to evidence regarding their expected benefits and harms, together with the patient’s previous treatment history and disease characteristics. |
| **Reopening / reconsideration** | What became reconsiderable was whether the population-derived evidentiary ordering should by itself determine which treatment possibilities remained clinically legitimate for this individual patient. The newly recommended treatment remained medically warranted by stronger evidence, but the limits of the available comparisons were also considered: population-level evidence did not directly compare all clinically possible treatment sequences, including every possibility of rechallenge. Rechallenge therefore remained a clinically possible option, although supported by more limited and uncertain evidence than the newly recommended treatment. |
| **Clinical response / provisional closure** | The patient came to understand the basis for her treating physician’s recommendation and chose to follow the recommended treatment. |

**Case 26**

| **Analytic component** | **Concise case-level account** |
| --- | --- |
| **Patient reality / consequential mismatch** | A patient had undergone surgery and chemotherapy for breast cancer and was subsequently advised to receive radiotherapy in accordance with standard treatment recommendations. She felt that she did not want any further treatment, but was uncertain whether she should nevertheless undergo radiotherapy because it was medically recommended. |
| **Existing medical framing** | Evidence-based medicine enables treatments to be recommended according to evidence of benefit and harm derived from populations, and such evidence is translated into clinical guidelines and standard treatment pathways. Within this framework, when an additional treatment has demonstrated benefit in reducing recurrence risk, completing that treatment becomes the medically recommended course for an eligible patient. |
| **Reopening / reconsideration** | What became reconsiderable was what authority a recommendation derived from population-level evidence should have in determining how the patient used her time and effort in her life. The clinician and patient considered both the medical grounds for recommending radiotherapy and what population-derived evidence could and could not determine for an individual patient. Against this uncertainty, they reconsidered whether reducing future recurrence risk should take priority over other ways in which she wished to use her time and effort, including pursuits through which she sought to develop herself. Radiotherapy remained medically recommended, while the place its preventive purpose should occupy within her life remained for her to determine. |
| **Clinical response / provisional closure** | After understanding why radiotherapy was medically recommended and the possible consequences of foregoing it, the patient chose not to undergo radiotherapy. |

**Case 27**

| **Analytic component** | **Concise case-level account** |
| --- | --- |
| **Patient reality / consequential mismatch** | A patient with breast cancer felt uneasy about relying solely on established cancer treatment and was interested in approaches outside conventional oncology. She wondered whether there might be something outside established medical treatment that could also help her. |
| **Existing medical framing** | Scientific medicine distinguishes clinically warranted interventions from those lacking sufficient evidence of efficacy and safety. This distinction is essential to responsible clinical practice. |
| **Reopening / reconsideration** | What became reconsiderable was what responsibility medicine could assume when asking a patient to entrust herself to treatment whose outcome for her could not be known in advance. This did not negate the established efficacy of evidence-based cancer treatment, but raised the question of whether that distinction alone was sufficient to respond to what the patient was seeking. The distinction remained medically necessary, while the clinician and patient considered why she was looking beyond medicine in the first place and what uncertainty remained unresolved even when medically warranted treatment was available. |
| **Clinical response / provisional closure** | After discussing what established cancer treatment could reliably offer and the limits of what it could provide, the patient chose to pursue both evidence-based cancer treatment and an approach without established anticancer efficacy, understanding the different evidentiary status of the two. |

**Case 28**

| **Analytic component** | **Concise case-level account** |
| --- | --- |
| **Patient reality / consequential mismatch** | The patient sought consultation because she felt unable to communicate what mattered to her within the relationship with her treating physician. Although her physician explained his recommendations in simple terms, she could not understand the reasoning through which those recommendations had been reached. Providing conclusions or treatment options alone therefore did not adequately address her difficulty. |
| **Existing medical framing** | Clinical medicine ordinarily requires physicians to define the clinical problem, determine what information and evidence are relevant, interpret uncertainty, weigh competing considerations, and formulate a recommendation. Much of this reasoning remains an internal professional process. |
| **Reopening / reconsideration** | What became reconsiderable was whether medicine’s own mode of reasoning could remain implicit when it was not intelligible to a patient without medical training. The clinician made explicit that medicine reasons from concepts, assumptions, and forms of knowledge that are not necessarily self-evident outside medicine, and explained from that foundation the medical issues that had been difficult for the patient to understand. The medical reasoning itself thereby became available for examination in dialogue with the patient. |
| **Clinical response / provisional closure** | The patient stated that she was able to understand the medical reasoning once its underlying concepts and assumptions had been made explicit. |

**Case 29**

| **Analytic component** | **Concise case-level account** |
| --- | --- |
| **Patient reality / consequential mismatch** | The patient had received different medical recommendations concerning different aspects of her health. She could understand the reasoning behind each one, but was uncertain what she should actually prioritize when they pointed in different directions. |
| **Existing medical framing** | Medicine makes complex bodily reality knowable by separating it into clinically tractable problems and generating knowledge about each of them. Each problem can therefore be investigated and managed through medically warranted forms of knowledge. |
| **Reopening / reconsideration** | Medicine’s capacity to reintegrate realities made knowable through reduction became reconsiderable. Separately warranted forms of medical knowledge did not by themselves yield an integrated judgment about what should take priority for the patient as a whole. The limits of such knowledge in resolving competing considerations were made explicit, allowing the patient’s own judgment to enter where medicine alone could not determine the priority. |
| **Clinical response / provisional closure** | The medical grounds underlying the different recommendations were made explicit, together with the limits of what each could determine when applied to the patient as a whole. The patient could then consider what she regarded as the appropriate priority for her own body while remaining informed by the relevant medical considerations. |

**Case 30**

| **Analytic component** | **Concise case-level account** |
| --- | --- |
| **Patient reality / consequential mismatch** | As one treatment followed another, the patient felt that something of her own life was being negated. Although she understood that the treatments were medically necessary, she struggled to understand what she had lived through as part of her own life. |
| **Existing medical framing** | Medicine organizes illness into a clinically coherent process through diagnosis, assessment of disease, treatment selection, evaluation of response, and subsequent management. Within this framing, illness and treatment can be understood as a sequence of medically meaningful events directed toward controlling disease and improving outcomes. |
| **Reopening / reconsideration** | What became reconsiderable was medicine’s capacity to respond to illness as a human experience. The clinician and patient considered how the reductionist organization of medical care into successive treatments, while medically warranted, could leave a person living through illness feeling overwhelmed or exhausted within the continuing sequence of care. The clinician acknowledged and shared the patient’s sense of exhaustion rather than treating it as a problem to be resolved by further medical intervention. |
| **Clinical response / provisional closure** | The patient was told that she could return to the clinic when she felt exhausted by the continuing course of treatment, and that the clinic could remain a place where such experiences could be brought and discussed. She accepted this understanding of the clinic’s role. |

**Case 31**

| **Analytic component** | **Concise case-level account** |
| --- | --- |
| **Patient reality / consequential mismatch** | A patient undergoing cancer treatment asked, “What about my life—how long might I live?” Although treatment had been effective, she remained uncertain about what this meant for her own future and how she should understand the life for which she was continuing treatment. |
| **Existing medical framing** | Medicine uses disease characteristics, treatment response, and population-derived evidence to estimate prognosis and evaluate treatment benefit. Within this framing, prolongation of survival is a central and measurable treatment outcome, even though the length of an individual patient’s life cannot be known in advance. |
| **Reopening / reconsideration** | What became reconsiderable was whether the value medicine sought to provide should be understood solely in terms of prolongation of life. The clinician and patient distinguished between medicine’s capacity to estimate and potentially extend the duration of life and its inability to determine how long this particular patient would live. They then considered what it meant for the patient to live within a future whose duration medicine could seek to extend but could not determine. |
| **Clinical response / provisional closure** | Prolongation of life could remain a meaningful aim of treatment without determining the value of the life for which treatment was being pursued. The clinician explained that her response to treatment provided grounds for expecting a longer life, while making clear that its actual duration could not be known. They discussed the importance of finding value in life beyond its length and of living fully within the life presently available to her. The patient said that she was able to accept this understanding. |
